# MSM with HIV: Improving prevalence and risk estimates by a Bayesian small area estimation modelling approach for public health service areas in the Netherlands

**DOI:** 10.1101/2022.05.20.22275273

**Authors:** Haoyi Wang, Chantal den Daas, Eline Op de Coul, Kai J Jonas

## Abstract

In many countries, HIV infections among MSM (MSMHIV) are closely monitored, and updated epidemiological reports are made available annually, yet the true prevalence of MSMHIV can be masked for areas with small population density or lack of data. Therefore, this study aimed to investigate the feasibility of small area estimation with a Bayesian approach to improve HIV surveillance. Data from the European MSM Internet Survey 2017 (EMIS-2017, Dutch subsample, n=3,459) and the Dutch survey ‘Men & Sexuality-2018’ (SMS-2018, n=5,653) were utilized in this study. We first applied a frequentist calculation to compare the observed relative risk of MSMHIV per Public Health Services (GGD) region in the Netherlands. We then applied a Bayesian spatial analysis and ecological regression to account for variance due to space and determinants associated with HIV among MSM to obtain more robust estimates. Results of the prevalence and risk estimations from EMIS-2017 and SMS-2018 converged with minor differences. Both estimations confirmed that the risk of MSMHIV is heterogenous across the Netherlands with some GGD regions, such as GGD Amsterdam [RR=1.21 (95% credible interval 1.05-1.38) by EMIS-2017; RR=1.39 (1.14-1.68) by SMS-2018], having a higher-than-average risk. Results from our ecological regression modelling revealed significant regional determinants which can impact on the risk for MSMHIV. In sum, our Bayesian approach to assess the risk of HIV among MSM was able to close data gaps and provide more robust prevalence and risk estimations. It is feasible and directly applicable for future HIV surveillance as a statistical adjustment tool.

## Introduction

### Background

For epidemiology in HIV, data are often characterised by a spatial or a spatio-temporal structure [1], however most studies in the field of HIV often ignore these spatial characteristics during data analysis. Examining the data provided by spatial analysis allows to identify men who have sex with men (MSM) with HIV (MSMHIV) clusters and to explore how these clusters originate [1, 2]. Therefore, spatial information and influence are likely to produce better surveillance models [1, 3, 4] in the context of a declining HIV epidemic in certain Global North regions, and reaching the 90-90-90 goals set by UNAIDS [5, 6].

For instance, in the Netherlands, despite annual epidemiological reports of MSMHIV having been provided by Stichting HIV Monitoring (SHM, the Dutch HIV monitoring foundation) [7], more accurate estimates on the spatial distribution are still desirable to close data gaps and to identify contexts in need of targeted interventions. Precise spatial distribution models of MSMHIV are crucial to assess areas of increased intervention need, to better programme services and to eventually end the HIV epidemic [8]. In line with the suggestion from Khan et al., we agree that HIV monitoring should go beyond urban/rural distinction to better inform policymakers [8], and spatial analysis on a smaller geographical scale is necessary. However, in the case of incomplete data for areas with small populations, the true risk of HIV can be hidden. Hence, more advanced techniques and methodologies are required to obtain robust estimates.

To respond, multiple studies proposed and used different small area estimation (SAE) techniques: From generalized additive models [9], over basic area-level models [10], to Poisson regression models [8]. In this study, taking the Netherlands as an example, we propose a Bayesian solution, which has been shown to be particularly effective and has been applied in several other epidemiological fields [3, 11-13], to estimate the posterior distribution (the revised or updated probability of an event occurring after considering new information and other uncertainties in the Bayesian inference [14]) of the prevalence and the risk of MSMHIV.

### Bayesian approach to estimate better HIV clusters

Comparing aforementioned approaches which usually fail to pick up the random effects due to fixed geographical foci, Bayesian modelling allows to account for similarities based on the neighbouring regions or by proximity, and to present data on a spatial hierarchical structure that borrows strength from the overall geo-spatial entity [3]. This hierarchical structure can thus be considered as a multi-level component which makes it possible to smoothen estimates based on the structural relationship during the structured random effects estimation instead of only treating the spatial information as a factor. Consequently, the smoothened HIV prevalence per small area by accounting for the geo-spatial structure can be more robust and closer to the true prevalence [3]. It would thus be possible to estimate a relative risk (RR) of HIV per area compared to the whole of the Netherlands, and to identify regions with higher-than-average risk of MSMHIV [3].

In addition, it is important to understand socio-ecological facilitators and hurdles of MSMHIV jointly with spatial distribution to unravel co-variations. A previous study has provided insights into the determinants of HIV transmission in the Netherlands on individual level, such as HIV testing, older age, and other diagnosed sexually transmitted infections (STIs) [15]. To evaluate these determinants of HIV transmission in the Netherlands from an ecological perspective to better understand the national and local MSMHIV epidemic, we also included these risk factors in a Bayesian spatial ecological regression model to explore how these risk factors impact on the HIV spatial distribution in the Netherlands (for more details see Methodology section and Online supplementary material).

This study thus sought to use data more effectively in order to generate opportunities to better understand the HIV epidemic by identifying the regions with higher-than-average risk of MSMHIV, using data from the Netherlands. We also aimed to fill the knowledge gap of how areal characteristics may impact MSMHIV by applying Bayesian spatial modelling methodology to provide a more accurate epidemiologic spatial pattern of MSMHIV using two independent survey-based datasets on HIV among MSM in the Netherlands. In addition, we compared the results by two survey-based datasets to explore the stability and robustness of Bayesian spatial analysis. Moreover, we compared the results with/without Bayesian inference to support future HIV surveillance and to support local HIV prevention efforts.

## Methodology

### Study population and data sources

#### EMIS-2017

All MSM included in this dataset were recruited between 19 October 2017 and 30 January 2018 via the European MSM Internet Survey (EMIS-2017, www.emis2017.eu) and were drawn from the Dutch subsample. EMIS-2017 was an anonymous, self-administered, and cross-sectional online survey conducted across 50 countries to inform interventions for MSM which are highly affected by infections with HIV and other STIs [16]. EMIS-2017 recruited 3,851 MSM in the Netherlands. We excluded 392 (10.2%) men that failed to provide information on their place of residence (final dataset n=3,459). Ethical approval for this survey was obtained from the Observational Research Ethics Committee at the London School of Hygiene & Tropical Medicine (review reference 14421 /RR/8805).

#### SMS-2018

The cross-sectional Survey Men & Sexuality (SMS-2018), led by Soa Aids Netherlands and Utrecht University, aimed to investigate the health, well-being, and sexuality of MSM in the Netherlands. It enrolled MSM between February and June 2018 through social media, gay media and dating apps. The survey was distributed in six languages to include a culturally diverse sample of MSM. All participants had provided informed consent prior to accessing the survey [17]. SMS-2018 recruited 6,206 MSM in the Netherlands. We excluded 552 (8.9%) men failed to provide their postal code (final dataset n=5,654). Ethical approval for this survey was obtained from the Ethics Committee of the Faculty of Social and Behavioural Sciences, Utrecht University (FETC17-131).

#### Study area

There are 25 Public Health Services (GGD) regions in the Netherlands. The GGDs are organized regionally and independently responsible to provide healthcare services and prevention, and to monitor the general health of the population. Therefore, by summarizing results on the GGD level, we can better support both local and national HIV monitoring and prevention. The geographical structure of the Netherlands with boundary geoinformation of GGD regions were retrieved from Statistics Netherlands (CBS) [18] and linked with postal code datasets [19].

All MSM data retrieved were deidentified and were aggregated on GGD regional level to represent HIV cases among MSM in the Netherlands separately from both datasets. Out of both samples, we included all MSM who provided information on 2-digit postal code (4-digit postal code for SMS-2018), which were then linked to the GGD regions. All data-aggregation processes were done separately for the two datasets without combination, hence there are no overlapping observations concerns in this study, given that participants could participate in both surveys.

### Small area estimation analysis

#### Frequentist analysis

We first calculated the observed HIV prevalence per GGD region with its 95% confidence interval (95%CI) by dividing the HIV counts by the numbers of MSM inhabitants participated in the surveys per region. We then calculated the observed standardized prevalence ratio (SPR) per GGD region, which is defined as the ratio of the observed counts to the expected counts using an indirect standardization approach, based on the overall risk of HIV in the Netherlands and the total MSM population in each GGD region [20]. As a spatial epidemiological measure, SPR could be applied to present the risk of HIV per GGD region compared to the overall risk of HIV in the Netherlands on the regional/populational level. In GGD regions where SPR>1, this denotes a higher risk of occurrence of HIV compared to the overall risk across the country, and GGD regions where SPR<1, this denotes a lower risk. However, in turn of regions with smaller populations sizes, SPR can be insufficiently reliable and stable due to pure chance calculation [21].

#### Bayesian modelling (null model)

To estimate a more accurate prevalence by Bayesian approach, we first conducted model accounting only the hierarchical spatial structure of GGD regions. Fig 1 presented the spatial structure on region connectivity of the Netherlands on GGD regional level based on the common sharing boarders or on proximity. To conduct the modelling analysis, we used the Integrated Nested Laplace Approximation (INLA), which is designed for latent Gaussian models, for the Bayesian computation. For the model parameters, we employed the Besag-York-Mollie (BYM) model, which specifies the spatially structured residual using an intrinsic conditional autoregressive (iCAR) distribution [22]. In addition, to specify the prior distribution for the Bayesian modelling, due to lack of information on GGD regions in the Netherlands, we assigned a weak, understandable, conservative and useful Penalized Complexity (PC) prior for the precision of the exchangeable random effects. For more detailed model assumptions and prior distribution, please see the online resource file S1.

**Fig 1.**
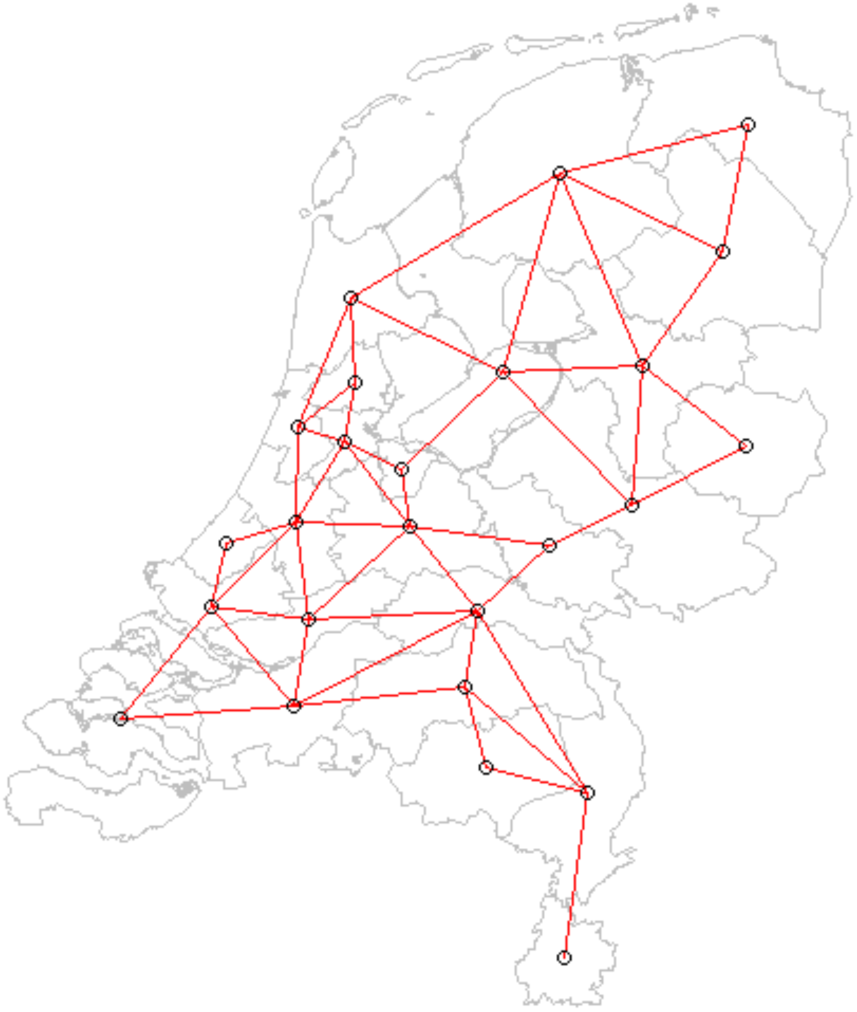
Region connectivity matrix of the Netherlands at the Public Health Services level. Note: for names and more details on the Public Health Services regions in the Netherlands, please see: https://www.ggd.nl/.

#### Bayesian spatial ecological regression modelling (final model)

Additional to the null model, we hypothesised that MSMHIV across the Netherlands can be influenced by the established determinants of MSMHIV reported by den Daas et al. [15] using EMIS-2010 datasets. These determinants included: prevalence of HIV testing (% ever tested); age (% >=35 years old); median number of sex partners; proportion of injecting drug users [(IDU), % IDU in EMIS-2017]/proportion of injecting drug use during sex [(SLAM), % SLAM in SMS-2018]; proportion of never using condom with last partner (% never); prevalence of syphilis (% yes), prevalence of chlamydia (% yes) and prevalence of gonorrhoea (% yes) in EMIS-2017 and prevalence of syphilis, chlamydia and gonorrhoea in the past 6 months in SMS-2018. Detailed definition of variables can be found in the published methodology paper for EMIS-2017 [16], and SMS-2018 [17].

Therefore, we applied a spatial ecological regression modelling technique [3] which takes into account these selected determinants of MSMHIV summarized by aggregating the selected datasets. We first conducted univariable models which only include one of the selected regional determinants and the spatial connectivity. We then conducted multivariable models with the significant determinants indicated by the univariable models to evaluate the impact on HIV prevalence in the Netherlands. We selected the final model using the backward approach by comparing models’ Deviance Information Criterion (DIC). The smaller DIC indicates the better goodness of fit in model. In other words, estimates by model with a smaller DIC are more robust, and can be consider closer to the true prevalence. For more detailed model assumptions and parameters, please see the online resource file S2. Finally, we quantified an Intra-class correlation (ICC) to evaluate the proportion of variance explained by the structured spatial component [3], and quantified the spatial random effects per GGD region based on the spatial structure of the Netherlands to estimate the influence from the spatial structure of the Netherlands on the GGD regional level on HIV prevalence for both the null spatial model and the final spatial ecological regression model.

### Computational analysis

All analyses were conducted in R (version 4.0.4). For all Bayesian modelling analyses with INLA, we used R-INLA package (version 21.05.02) to empower our computational process [3].

## Results

In this section, we first present the GGD regional characteristics and the results of the frequentist analysis to show the spatial distribution of HIV in the Netherlands by GGD regions, using a classical frequentist approach for both datasets. We then present the posterior prevalence and RR of HIV per GGD region estimated by Bayesian modelling with only taking the spatial structure into account (null model). By comparing the results from the null model and the naïve analysis, we can identify how the spatial structure itself impacts on the distribution of HIV in the Netherlands beyond the chance levels. Next, we include the procedure of model selection by presenting the univariable and multivariable regression modelling results to unravel which known individual-level determinants of HIV are significantly related to HIV status on the areal level. Finally, the posterior prevalence and RR of HIV per GGD region, estimated by the multivariable regression modelling (final model), are presented to be compared with the results from the null model. The comparison can, therefore, inform how the HIV spatial distribution could change after conditioning other determinants of HIV.

### Study population characteristics

Regional characteristics relevant to MSMHIV across the Netherlands were heterogenous for both datasets (see Online resource S1 table). For these established individual level determinants, HIV testing proportions ranged from 64.1% in GGD Drenthe to 92.3% in GGD Amsterdam in EMIS-2017 (and ranged from 62.7% in GGD Limburg-Noord to 92.7% in GGD Amsterdam in SMS-2018). Major differences between ever-/recent-diagnosed STIs proportion among Dutch MSM were observed from the two datasets. For ever diagnosed STIs proportion in EMIS-2017, ever diagnosed syphilis ranged from 10.3% in GGD Drenthe to 34.4% in GGD Zaanstreek/Waterland; and ever diagnosed chlamydia ranged from 19.7% in GGD Gelderland-Zuid to 45.2% in GGD Zaanstreek/Waterland. For recent diagnosed STIs (within six months) in SMS-2018, recently diagnosed syphilis ranged from 18.0% in GGD Drenthe to 44.4% in GGD Amsterdam. More detailed information for other regional characteristics (older than 35 years proportion, IDU proportion, and other STIs) per GGD region from both datasets can be found in Online resource S1 table.

### Frequentist observed HIV prevalence and risk among MSM

In terms of the prevalence of MSMHIV in the Netherlands, the observed overall prevalence of HIV among MSM in 2017 was 14.2% in EMIS-2017 and 9.5% in SMS-2018. In EMIS-2017, the observed prevalence of HIV varied by GGD regions in the Netherlands, with a range of 6.8% (95%CI 3.16-14.09) in GGD Limburg Noord to 25.0% (95%CI 13.25-42.11) in GGD Zaanstreek/Waterland. In SMS-2018, the observed prevalence varied from 3.7% (95%CI 1.59-8.38) in Veiligheids-en Gezondheidsregio Gelderland-Midden (VGGM) to 14.15% (95%CI 11.67-17.06) in GGD Amsterdam (Fig 2a&2d, Online resource S2 table). The crude SPR in Fig 3-a and 3-d shows that regions with higher-than-average risk of HIV exist in the Netherlands, with a range of 0.43 (GGD Limburg Noord) to 1.59 (GGD Zaanstreek/Waterland) in EMIS-2017; and 0.39 (VGGM) to 1.49 (GGD Amsterdam) in SMS-2018. The SPR trends corresponded with the patterns of the observed HIV prevalence. More detailed information for the frequentist observed prevalence and SPR of MSMHIV can be found in Online resource S2 table.

**Fig 2.**
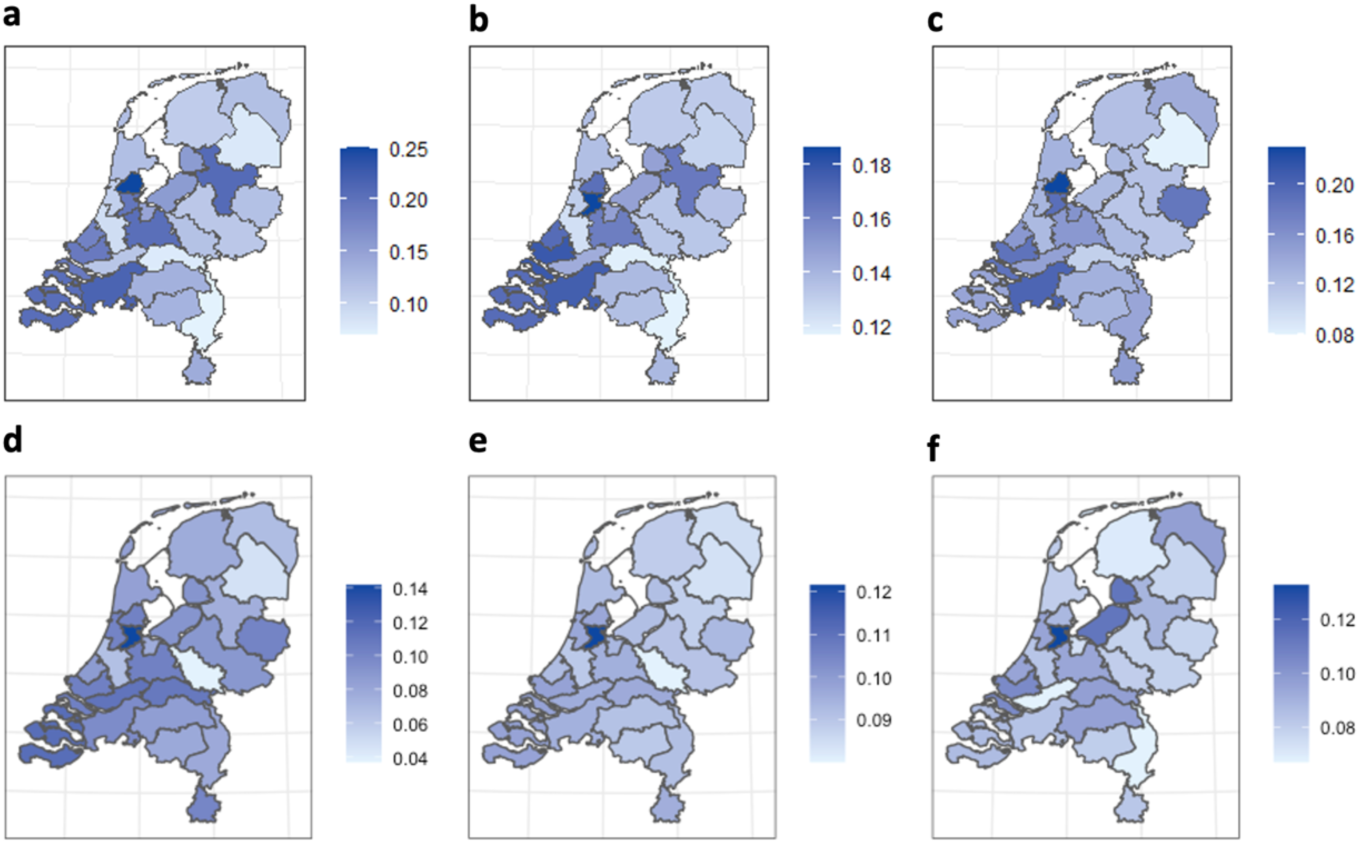
Choropleth map of the estimates of HIV prevalence by GGD regions in the Netherlands. A: Observed HIV prevalence by EMIS-2017. B: Posterior mean of HIV prevalence estimated by Bayesian spatial modelling (null model) by EMIS-2017. C: Posterior mean of HIV prevalence estimated by Bayesian spatial ecological regression modelling (final model) by EMIS-2017. D: Observed HIV prevalence by SMS-2018. E: Posterior mean of HIV prevalence estimated by Bayesian spatial modelling (null model) by SMS-2018. F: Posterior mean of HIV prevalence estimated by Bayesian spatial ecological regression modelling (final model) by SMS-2018. See Online resource table S2 for the 95%CI or 95%CrI and other details. Notes: the darker a GGD region, the higher the prevalence estimation of HIV among MSM.

**Fig 3.**
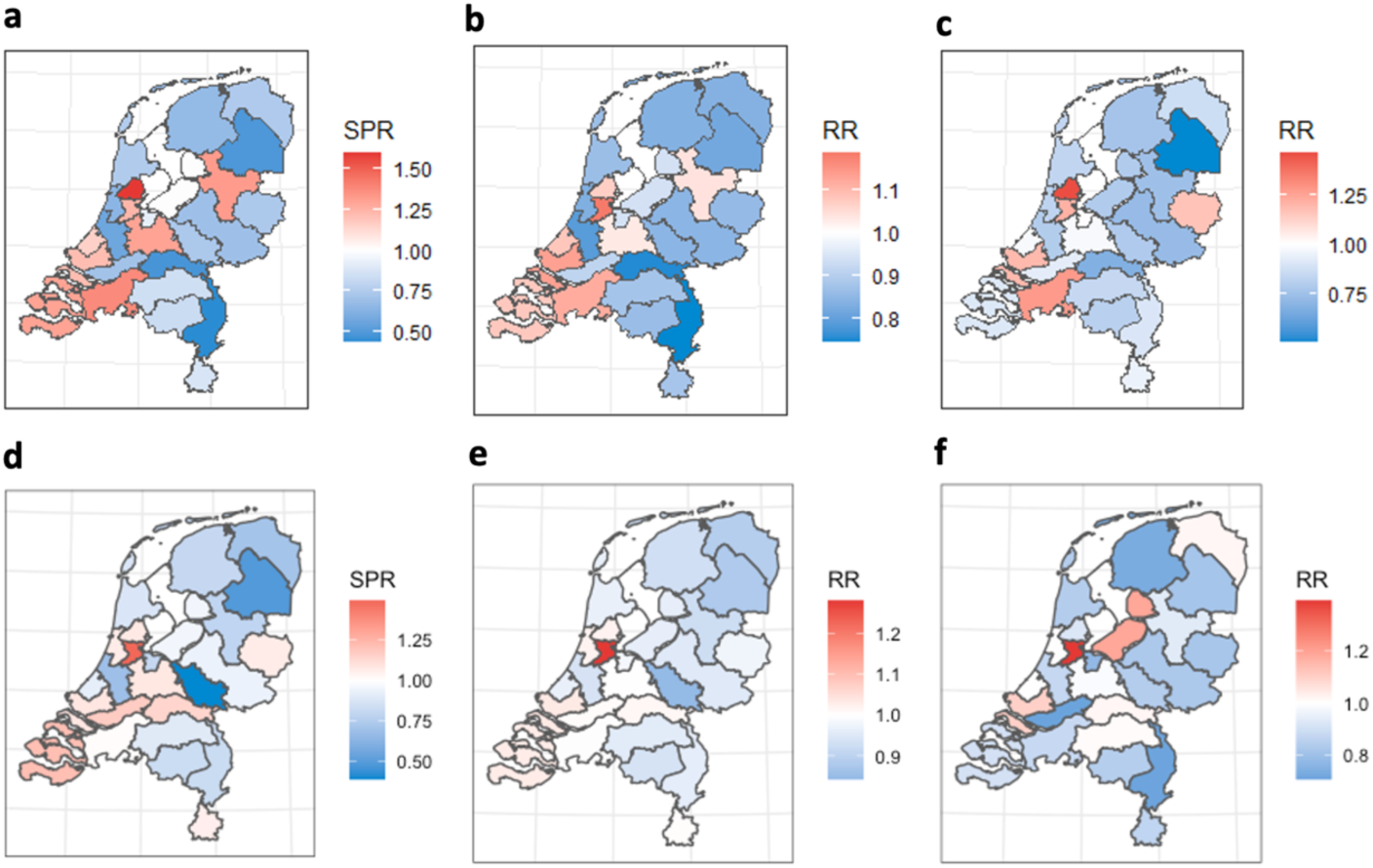
Choropleth map of the estimates of HIV risks by GGD regions in the Netherlands. A: Observed HIV standardised prevalence ratio by EMIS-2017. B: Posterior mean of HIV relative risk estimated by the Bayesian spatial modelling (null model) by EMIS-2017. C: Posterior mean of HIV relative risk estimated by the Bayesian spatial ecological regression modelling (final model) by EMIS-2017. D: Observed HIV standardised prevalence ratio by SMS-2018. E: Posterior mean of HIV relative risk estimated by the Bayesian spatial modelling (null model) by SMS-2018. F: Posterior mean of HIV relative risk estimated by the Bayesian spatial ecological regression modelling (final model) by SMS-2018. See Online resource table S2 for the 95%CI or 95%CrI and other details. Notes: RR (or SPR) higher than 1 indicates a higher-than-average (average risk in the Netherlands) risk of HIV among MSM in that region (red); RR (or SPR) lower than 1 indicates a lower-than-average risk of HIV among MSM in that region (blue).

### HIV prevalence and risk among MSM after Bayesian spatial adjustment

After accounting for the spatial effects based on the spatial structure of the Netherlands on the GGD region level presented in Fig 1 without other regional determinants of HIV transmission, the EMIS-2017 ICC of the spatial structure was estimated at 0.24, which indicates that around 24% of the observed variance of HIV among MSM in the Netherlands can be explained by the spatial structure of the Netherlands on the GGD regional level, and the SMS-2018 ICC was 0.27 (Table 1).

**Table 1.** Model comparison and selection for EMIS-2017 and SMS-2018.

| Models | HIV diagnosis |  |  |  |  |  |  |  |  |  |
| --- | --- | --- | --- | --- | --- | --- | --- | --- | --- | --- |
|  | EMIS-2017 |  |  |  |  | SMS-2018 |  |  |  |  |
|  | Covariates | Coefficient | 95%CrI | DIC | ICC | Covariates | Coefficient | 95%CrI | DIC | ICC |
| Spatial Null model | Intercept | -0.108 | (-0.257 – 0.025) | 145.71 | 0.24 | Intercept | -0.041 | ( -0.196 – 0.098 ) | 123.99 | 0.27 |
| Spatial Univariable models | Intercept | -2.359 | (-4.052 - -0.597) | 144.42 | 0.27 | Intercept | -2.206 | ( -3.729 - -2.217 ) | 118.97 | 0.31 |
|  | HIV test (%yes)* | 2.724 | (0.611 – 4.725) |  |  | HIV test (%yes)* | 2.674 | ( 0.738 – 4.501 ) |  |  |
|  | Intercept | -0.591 | (-1.376 – 0.172) | 145.89 | 0.24 | Intercept | -1.166 | ( -2.027 - -0.271 ) | 119.84 | 0.28 |
|  | Age (%>= 35 y.o.) | 1.089 | (-0.613 – 2.786) |  |  | Age (%>= 35 y.o.)* | 2.454 | (0.551 – 4.229) |  |  |
|  | Intercept | -0.063 | (-0.232 – 0.083) | 147.26 | 0.26 | Intercept | 0.073 | ( -0.498 – 0.645 ) | 125.66 | 0.27 |
|  | Partner | -0.037 | (-0.104 – 0.032) |  |  | Partner | -0.049 | ( -0.297 – 0.186 ) |  |  |
|  | Intercept | -5.305 | (-10.373 – 0.715) | 144.51 | 0.25 | Intercept | -0.971 | ( -1.763 - -0.130 ) | 121.04 | 0.29 |
|  | Condom (%never) | 41.315 | (-6.485 – 81.421) |  |  | Condom (%never)* | 3.121 | ( 0.359 – 5.622 ) |  |  |
|  | Intercept | -0.398 | (-0.752 - -0.056) | 145.51 | 0.32 | Intercept | 0.005 | ( -0.235 – 0.227 ) | 125.47 | 0.27 |
|  | IDU (%yes) | 4.41 | (-0.369 – 9.051) |  |  | SLAM (%yes) | -0.561 | ( -2.812 – 1.601 ) |  |  |
|  | Intercept | -0.87 | (-1.384 - -0.352) | 142.41 | 0.28 | Intercept | -0.049 | ( -0.327 – 0.220 ) | 125.62 | 0.27 |
|  | Syphilis (%yes)* | 3.546 | (1.263 – 5.703) |  |  | Syphilis (%yes)# | 0.251 | ( -8.189 – 8.139 ) |  |  |
|  | Intercept | -0.662 | (-1.281 - -0.029) | 146.6 | 0.24 | Intercept | -0.197 | ( -0.651 – 0.248 ) | 125.42 | 0.29 |
|  | Chlamydia (%yes) | 1.724 | (-0.200 – 3.514) |  |  | Chlamydia (%yes)# | 1.992 | ( -3.513 – 7.301 ) |  |  |
|  | Intercept | -0.856 | (-1.552 - -0.132) | 145.8 | 0.26 | Intercept | -0.125 | ( -0.533 – 0.277 ) | 125.64 | 0.27 |
|  | Gonorrhoea (%yes)* | 2.255 | (0.116 – 4.221) |  |  | Gonorrhoea (%yes)# | 1.195 | ( -4.376 – 6.498 ) |  |  |
| Spatial Multivariable final model | Intercept | -2.021 | (-3.624 - -0.358) |  |  | Intercept | -2.225 | (-3.701 - -0.739) |  |  |
|  | HIV test (%yes) | 1.607 | (-0.6 – 3.737) | 141.74 | 0.28 | HIV test (%yes) | 1.801 | (-0.286 – 3.941) | 118.58 | 0.30 |
|  | Syphilis (%yes) | 2.674 | (0.192 – 5.099) |  |  | Age (%>= 35 y.o.) | 1.515 | (-0.549 – 3.571) |  |  |

Note: \* = significant areal determinants of the univariable models. Partner = median number of partners. Condom = % never used condom with non-steady partners. # Indicates six-month prevalence instead of life-time prevalence. CrI = credible interval. DIC=Deviance Information Criterion, ICC= Intra-class correlation

As indicated by the observed HIV prevalence, we observed heterogeneity of the posterior HIV prevalence in the Netherlands estimated by the spatial null models in both datasets. In EMIS-2017, the highest posterior HIV prevalence was found in the GGD Amsterdam of 18.6% (95%CrI 15.87-21.58) and the lowest in GGD Limburg-Noord of 11.7% (95%CrI 7.2-16.4). GGD Amsterdam was estimated as the only region with statistically significant higher-than-average risk of HIV among MSM in the Netherlands with a RR of 1.18 (95%CrI 1.01-1.37). Full details of all GGD regions according to the spatial null model can be found in Online resource table S2 and Fig 3-b. In SMS-2018, the posterior prevalence and RR was the similar as the observed prevalence, the highest posterior HIV prevalence was found in Amsterdam, too, of 12.16% (95%CrI 9.58-15.14) with the only significant higher-than-average risk of HIV of 1.28 (95%CrI 1.01-1.59), and the lowest in VGGM of 8% (95%CrI 5.09-10.69) with RR of 0.84 (95%CrI 0.54-1.13), see Fig 3-e and Online resource S2 table.

Posterior spatial random effects on the HIV prevalence estimated by the null model can be found in Fig 4-a and 4-c, which confirms the spatial heterogeneity and indicates how the spatial structure impacts on the estimated posterior RR per GGD region, with a range of [EMIS-2017: −0.21 (GGD Limburg-Noord) to 0.27 (GGD Amsterdam)] and [SMS-2018: −0.15 (VGGM) to 0.28 (GGD Amsterdam)]. In other words, regions with a positive (or negative) value of the spatial random effects indicate having an elevated (or lower) relative risk of HIV than the overall risk in the Netherlands.

**Fig 4.**
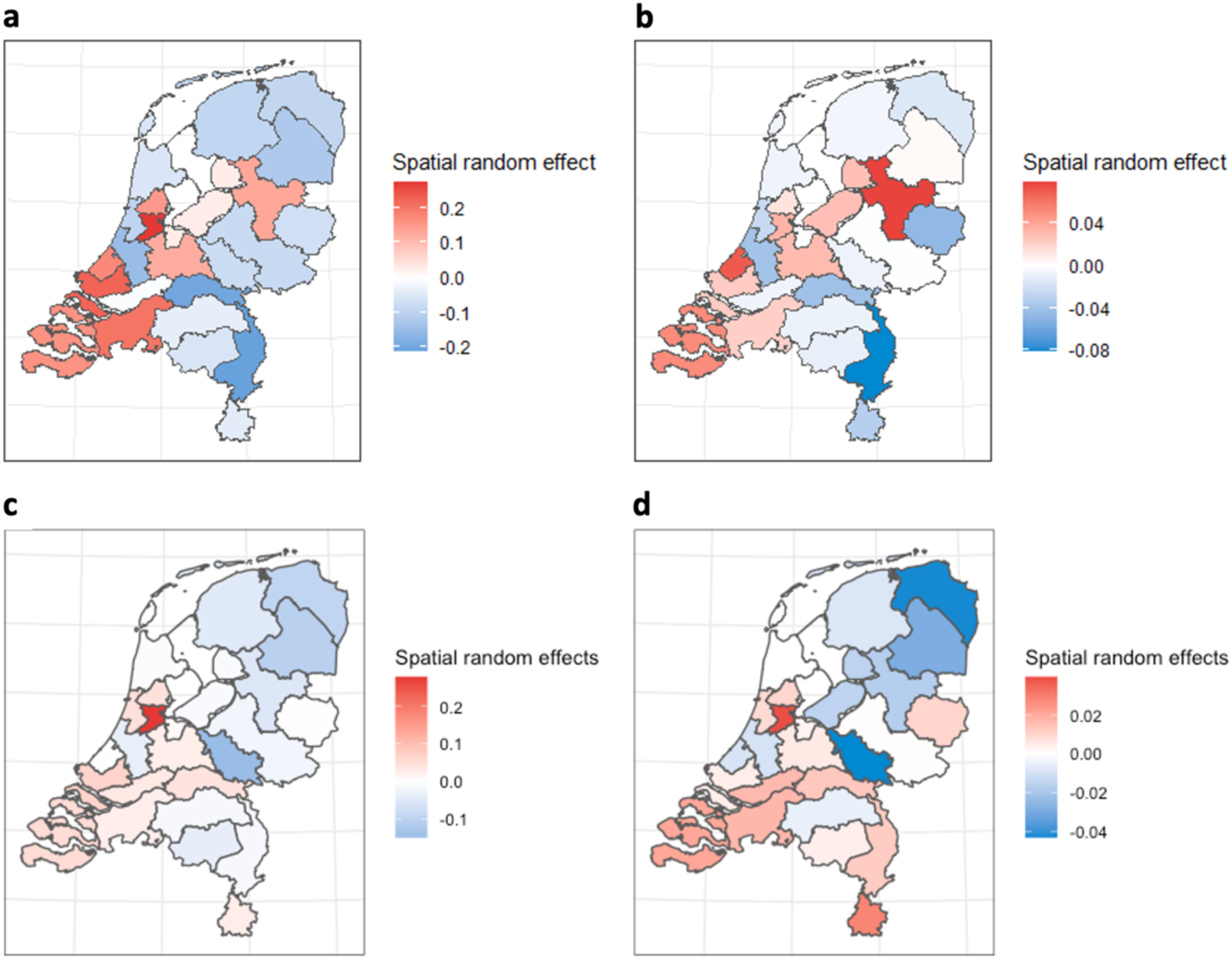
Posterior spatial random effects on the GGD regional level in the Netherlands. A: posterior spatial random effects estimated by Bayesian spatial modelling (null model) by EMIS-2017. B: Posterior spatial random effects estimated by the Bayesian spatial ecological regression modelling (final model) by EMIS-2017. C: posterior spatial random effects estimated by Bayesian spatial modelling (null model) by SMS-2018. D: Posterior spatial random effects estimated by the Bayesian spatial ecological regression modelling (final model) by SMS-2018. See Online resource table S2 for the 95%CrI and other details. Notes: regions with a positive (negative) value of the spatial random effects indicate having an elevated (lower) relative risk of HIV among MSM than the overall risk in the Netherlands.

### HIV prevalence and risk among MSM after Bayesian spatial ecological adjustment

#### Univariable models

In EMIS-2017, after adjusting on the observed HIV testing prevalence as the regional determinant, a coefficient of 2.72 (95%CrI 0.61-4.73, DIC=144.42, ICC=0.27) was modelled. This means that each increase of one percent in HIV testing prevalence in a region is associated with an increase of around 2.8% (=exp(2.724*0.01)) in HIV risk in that region. The coefficient for the observed syphilis prevalence was estimated at 3.55 (95%CrI 1.26-5.70, DIC=142.41, ICC=0.28), which indicated that for every one percent increase of the regional prevalence of syphilis, the regional risk of HIV increases by 3.5%. Similar for gonorrhoea, the increased risk was 2.3%. The other areal determinants of HIV (age, number of partners, condom use, IDU prevalence and chlamydia prevalence were not significantly associated with the posterior mean of RR of HIV in the Netherlands (Table 1).

In SMS-2018, we observed a significant positive association between HIV test and HIV with a coefficient of 2.67 (95%CrI 0.74-4.50, ICC=0.31, DIC=118.97) in the univariable model. In addition, instead of STIs’ spatial prevalence, we found that proportion of higher age, coefficient=2.45 (95%CrI 0.55-4.23, ICC=0.28, DIC=119.84) and proportion of never using a condom with non-steady partners, coefficient=3.12 (95%CrI 0.36-5.62, ICC=0.29, DIC=121.04) were positively associated with the HIV prevalence on the GGD regional level. Other areal characteristics of GGD regions were not estimated significant within this dataset (Table 1).

#### Multivariable models (final model)

After conditioning significant areal determinants of HIV, and selecting by the smallest DIC, in EMIS-2017, the final model included HIV testing prevalence with a coefficient of 1.60 (95%CrI −0.60-3.74) and syphilis prevalence with a coefficient of 2.67 (95%CrI 0.19-5.10), a DIC of 141.74, and an ICC of 0.28. The coefficients’ estimations indicate that both univariable models of HIV testing, and ever-diagnosed syphilis prevalence overestimated the effects from these two regional determinants of HIV. Even though HIV testing was not statistically significant in the final model, the DIC of the final model was smaller than the DIC of the model with only syphilis prevalence (DIC=142.41). Therefore, we kept HIV testing in the final model (Table 1). In other words, with a smaller DIC, the posterior prevalence and RR of MSMHIV estimated should be closer to the true prevalence and RR. In the multivariable model based on SMS-2018, we included HIV test prevalence with coefficient=1.80 (95%CrI −0.29-3.94) and proportion of higher age with coefficient=1.515 (95%CrI −0.55-3.57) with the smallest DIC of 118.58 and an ICC of 0.30 in the final model (Table 1).

The posterior prevalence of HIV was again heterogenous with both datasets in the Netherlands. In EMIS-2017, the highest posterior prevalence of HIV was observed from GGD Zaanstreek/Waterland of 22.9% (95%CrI 16.25-30.8), and the lowest posterior prevalence of HIV was observed from GGD Drenthe of 7.89% (95%CrI 4.94-12.02). In addition, the final model succeeded to pick up the regions with higher-than-average risk of MSMHIV in the Netherlands other than GGD Amsterdam (RR=1.21, 95%CrI 1.05-1.38): GGD Rotterdam-Rijnmond (RR=1.19, 95%CrI 1.00-1.41) and GGD Zaanstreek/Waterland (RR=1.46, 95%CrI 1.04-1.96). Also, the risk of HIV of GGD Noord-en Oost-Gelderland (RR=0.72, 95%CrI 0.54-0.94), GGD Fryslân (RR=0.76, 95%CrI 0.57-0.98), GGD Drenthe (RR=0.5, 95%CrI 0.31-0.77) and GGD Hollands-Midden (RR=0.79, 95%CrI 0.58-0.99) were found significantly lower than the risk on the average level in the Netherlands. In SMS-2018, the lowest posterior prevalence was estimated in Dienst Gezondheid & Jeugd ZHZ of 6.71% (95%CrI 4.69-9.47) with a RR of 0.71 (95%CrI 0.49-1.00) and GGD Limburg-Noord of 6.71% (95%CrI 3.46-11.42) with a RR of 0.71 (95%CrI 0.36-1.20). The highest posterior prevalence was again observed in GGD Amsterdam: 13.25% (95%CrI 10.84-15.93), and it was the only one GGD region with a significant higher-than-average risk of HIV in the Netherlands (RR=1.39, 95%CrI 1.14-1.68). For full details of posterior HIV prevalence and RR per GGD region, see Online resource table S2 and Fig 3-c and 3-f.

The spatial random effects indicated again the heterogeneity on the RR of MSMHIV across the Netherlands with a range of [EMIS-2017: −0.08 (GGD Limburg Noord) to 0.08 (GGD Ijsselland); SMS-2018: −0.04 (VGGM) to 0.04 (GGD Amsterdam)]. The posterior spatial random effect per GGD region were summarised in Online resource table S2 and Fig 4-b and 4-d.

## Discussion

To illustrate the usefulness of SAE modelling with a Bayesian approach, we investigated the spatial distribution of MSMHIV in conjunction with determinants of MSMHIV using data from the Netherlands at the level of the Public Health Services regions (GGD). We applied this methodology on two independent survey-based datasets to explore the applicability and the estimates of accuracy for the HIV surveillance at the same time.

Based on both datasets, we observed a heterogenous spatial distribution of MSMHIV: There are GGD regions which showed higher-than-average risk. In particular, the GGD Amsterdam region as expected, and GGD Zeeland, had a significantly higher-than-average risk of MSMHIV in the Netherlands. Jointly with the spatial patterns, we identified regional determinants to be significantly associated with MSMHIV prevalence in the Netherlands. Methodologically, we found that the observed prevalence estimated by the frequentist analysis was less stable than the posterior prevalence estimated by the Bayesian spatial modelling in terms of the estimations range and their uncertainty range (the 95% confidence interval and 95% credible interval), especially for regions with smaller sample sizes. Despite a largely overlapping spatial distribution and heterogeneity of HIV in the Netherlands (both Frequentist observed, and Bayesian smoothed) between the two datasets, minor differences in the prevalence and spatial random effects were obtained.

### Spatial distribution of HIV among MSM in the Netherlands

Overall, based on the overlapping results from both datasets, we observed a higher prevalence of HIV in the West of the Netherlands where also the main urban areas (in Dutch: Randstad) are located, and in the GGD region of Zeeland, which belongs to the area that has the highest concentration of conservative orthodox Calvinist Protestants in the country [23].

It was within our expectation that the prevalence of MSMHIV was higher in the GGD regions in the Randstad, such as GGD Amsterdam. This prevalence is also in line with findings from previous studies using surveillance data by geographic information system and survey-based data [15, 24]. In addition, our analysis based on both datasets suggested a significant higher-than-average risk of HIV among MSM in this GGD region compared to other regions in the Netherlands. Few reasons may explain our findings. Firstly, Amsterdam which is known as the ‘Gay Capital of Europe’ is the target of “gay tourism”, with more sexual encounters occurring subsequently. [25]. Likewise, more Dutch MSM choose to relocate to these main urban areas [17], and the HIV cases would, therefore, be concentrated there as well. Another reason that contributed to a higher HIV prevalence is the high HIV testing rate among MSM in GGD Amsterdam region (Online resource S1 table). Our ecological modelling analysis also confirmed this argument that with a higher HIV testing prevalence, the risk of HIV in that region would be higher as well (Table 1).

It was, however, not expected that the GGD Zeeland also had a higher spatial risk (random effect, Fig 4) of HIV among MSM compared to other regions. One reason for this higher risk found for this region may be due to the religion/local culture. As one of the most conservative regions in the country with associated negative views on same-sex sexual activities and relations, an overall negative attitude towards homosexuality may be greater than in other regions [26]. In turn, some sexual behaviours may be stigmatized and MSM may experience more barriers to HIV testing, which may influence the risk of MSMHIV at that region: according to both datasets GGD Zeeland has one of the lowest HIV test prevalence among MSM (Online resource S1 table). Second, a longer distance to the STI clinics could play a role as a barrier to HIV testing in GGD Zeeland [27], which could also leave an influence on the spatial distribution of MSMHIV in the Netherlands. Therefore, future studies should also investigate the distance to the STI clinics as a regional determinant for a more comprehensive model.

### Differences between estimations by EMIS-2017 and SMS-2018

Despite the large overlap of the spatial pattern of MSMHIV by our analysis based on EMIS-2017 and SMS-2018 datasets, we observed some minor differences in terms of both the observed and estimated posterior prevalence, which is generally lower in SMS-2018 data compared to EMIS-2017 data. One reason that may explain this finding is that data collection variations existed between EMIS-2017 and SMS-2018. The collection methods and process were different between the SMS-2018 and EMIS-2017. It would thus lead to collection variations and resulted in different estimates. However, in terms of the posterior relative risk of HIV on the GGD regional level, the results between these two datasets converged and can reflect how the regional risks differed between different GGD regions on the national scale. This converged posterior RR estimations by both datasets indicate the strong stability of the Bayesian spatial analysis to identify regions with higher risk for prevention efforts allocation strategies.

We also observed a different impact of the areal determinants on our HIV prevalence and risk modelling between these two datasets. Despite the discussed sampling variations, different definitions of the determinants when collecting data through the surveys could also explain why our univariable models and final models are different. For example, in EMIS-2017, men were asked if they were ever diagnosed with any type of STI instead of STI diagnosis within the past six months, as in SMS-2018. The HIV-risk profile and sexual behaviour profile of a MSM would thus be different and result in different impact on the ecological modelling analysis. Therefore, based on our findings in the univariable models, we could also conclude that the impact of the lifetime STI diagnoses should be greater than the recent STI diagnoses.

### Application of Bayesian spatial modelling analysis

The application of Bayesian spatial modelling analysis in two survey-based datasets from the Netherlands proved that modelling the HIV distribution with a Bayesian approach is feasible, and robust when comparing results between two datasets. Compared to calculating the observed prevalence and SPR, results of the posterior prevalence and risks estimations were smoother and more stable due to the narrower credible intervals estimated by INLA, which has been proven helpful to estimate the more accurate prevalence and risk as an approximation approach [3]. It thus delivers more certainty when interpreting the results and tailoring prevention programming for HIV.

In addition, our spatial ecological modelling allowed us to investigate the variations of HIV based on the spatial connectivity together with other regional determinants of HIV. We found several regional determinants (Table 1) based on our survey data useful to estimate the posterior prevalence and risk of HIV in the Netherlands. Consequently, both two final models for EMIS-2017 and SMS-2018 data improved the goodness of fit after adding the regional determinants related to MSMHIV, and we believe the estimations from the final model should be closer to the true prevalence compared to the observed frequentist calculation and models without covariates. Therefore, the established spatial determinants from this study should be considered valuable for policymakers and HIV surveillance authorities. Attention should be also given to these regional HIV determinants instead of focussing only the numerical prevalence only.

We thus recommend promoting this novel methodology as a statistical adjustment for future HIV national/local surveillance, especially when there are gaps due to missing data, or regional prevalence estimates are needed. Even though we acknowledge that the complex statistical computation, unfamiliarity and limited knowledge on Bayes’ Theorem may limit the application of this methodology for non-Bayesian stakeholders, the already available techniques and the various forms of open source statistical software [3, 28, 29] should help to ease the computation process and help interpreting results. We believe applying SAE with a Bayesian approach can help to robustly tailor HIV prevention programmes, especially local HIV prevention resources and services navigation.

### Strength and limitations

We acknowledge the following strengths and limitations of our study. One major strength of this study is the introduction and the application of Bayesian spatial modelling analysis as a SAE method. We considered our results, especially the posterior risks of MSMHIV, as robust and valuable for HIV related public health policies and prevention strategies. The methodology in our study can be directly applied in other countries in the future for small area estimations using surveillance data on HIV. Another strength is the convergence of the models based on data from two survey-based datasets. Data from these two surveys made MSM individual level covariates directly available for the posterior modelling analysis instead of using secondary area-level covariate data based on the Dutch general population. Moreover, presenting data on the GGD regional level also helped to prevent information bias due to the municipal location of HIV testing. Since the regional public health service runs the majority of HIV tests in the Netherlands and since the sexual health clinics are located in the larger municipalities in a GGD region, data may thus concentrate in these bigger cities if HIV among MSM would be assessed on the municipality level.

In addition to the aforementioned limitations, one limitation can be the lack of data from the neighbouring regions from other countries. Our Bayesian spatial analysis with a hierarchical structure revealed how regions may influence each other to smoothen the risk estimates based on neighbouring information or on proximity. However, given the smoothing by neighbouring regions, our analysis may be influenced by other regions outside the Netherlands. It should be stressed that for some GGD regions which are located in the border regions of the Netherlands, the estimated prevalence and risks of HIV of these regions would thus be less stable compared to the rest due to the lower predictability as only one other node is available and thus part of spatial information is missing. Regions that share a boarder with Germany and Belgium, especially for GGD Zuid-Limburg which is only geographically connected with GGD Limburg-Noord and without other neighbouring regions in the Netherlands (Fig 1), require additional cross-border data input. Therefore, a study including those neighbouring regions in Belgium and Germany may be warranted in the future to compensate for the problem of lack of national spatial connectivity for those boarder regions. To achieve this aim, comparable cross-border data needs to be accessible, too. Moreover, our spatial analysis of MSMHIV across the Netherlands was based on survey data from 2017-2018 when the pre-exposure prophylaxis (PrEP) has yet to be formally introduced in the Netherlands (2019). Our spatial model, therefore, did not include PrEP use among MSM per GGD region as a regional characteristic. Consequently, the influence from PrEP use was not measured in our models. Given the established impacted on the HIV prevention among MSM from using PrEP [30-34], future studies should therefore include PrEP use into the spatial models for a more robust estimation. Another limitation can be the lack of an informative prior distribution when conducting Bayesian spatial analysis. Previous studies which applied Bayesian statistic in other epidemiologic field has suggested that to acquire the true prevalence and RR, an informative prior is preferred and required in practice [35, 36]. Our application of the PC prior, as a weakly informative prior, may thus limit the robustness of our posterior estimation and make them conservative [36]. However, we believe our estimations were still robust and close to the true risk of HIV among MSM based on the previous sensitivity analysis of PC in a prior experiment [37]. In addition, even though our Bayesian approach made our estimations more robust, more comprehensive datasets, such as routine surveillance data are still warranted. Another major limitation in our study may be the lack of temporal dimension in our models. The scope of our study to offer a time-dynamic epidemiologic picture on how MSMHIV spatially distribute over the time is limited. Future studies thus should include a wide temporal period to support a more comprehensive spatio-temporal analysis. Finally, ecological fallacy is possible due to our ecological study design. We lose information on the individual-level due to aggregating information spatially. Our results on the roles of the regional characteristics thus cannot be directly applied to investigate/predict the MSM’s HIV risk profile on the individual level.

## Conclusion

In conclusion, our study proposed a Bayesian approach to more accurately assess the risk of HIV among MSM using data from the Netherlands on the public health service regional level with more robust prevalence and risk estimations over the use of crude proportions. Our findings based on two independent surveys can be considered valuable for policymakers and HIV surveillance authorities for resources allocation decision by prioritizing resources to the regions which require more efforts to reduce the burden of HIV among MSM accordingly. Based on the Dutch data, our method has shown to be feasible and can be directly applied to achieve a more comprehensive and robust surveillance of HIV in any geographic context.

## Data Availability

All data produced in the present study are available upon reasonable request to the authors.

## Acknowledgement

We thank all participants of the surveys and the individuals involved in preparation, execution and analysis of the surveys. We thank Wim Zuilhof, Bouko Bakker, Aryanti Radyowijati, Koenraad Vermey, Arjan van Bijnen and the EMIS board for their invaluable help for the EMIS-2017 data. In addition, we thank John de Wit, Philippe Adam, and Wim Zuilhof, for their role in the development and collection of SMS-2018 data.

## Statements & Declarations

### Author Contributions

All authors contributed to the study conception and design. Material preparation, data analysis were performed by [Kai J Jonas], [Chantal den Daas], and [Haoyi Wang]. The first draft of the manuscript was written by [Haoyi Wang] and all authors commented on previous versions of the manuscript. All authors read and approved the final manuscript.

### Ethics approval

This study was performed in line with the principles of the Declaration of Helsinki. Approval was granted by the Observational Research Ethics Committee at the London School of Hygiene & Tropical Medicine (review reference 14421/RR/8805) and the Ethics Committee of the Faculty of Social and Behavioural Sciences, Utrecht University (FETC17-131).

### Consent to participate

Informed consent was obtained from all individual participants included in the studies.

